# COVID-19 vaccination in California: Are we equitable yet?

**DOI:** 10.1101/2021.05.25.21257807

**Authors:** Alec J. Schmidt, Maria L. Daza-Torres, Yury E. García, Joan L. Ashby, Bradley H. Pollock, James Sharpnack, Miriam Nuño

## Abstract

**Background:** By March 2021, California had one of the least equitable COVID-19 vaccine distribution programs in the US. To rectify this, Governor Newsom ordered 4 million vaccine doses be reserved for the census tracts in the lowest quartile of the Healthy Places Index (HPI). California plans to lift state-wide restrictions on June 15th, 2021, as long as test positivity and vaccine equity thresholds are met in the state’s most vulnerable neighborhoods. This short investigation examines current vaccine equity and forecasts where California can expect to be when the economy fully reopens.

**Methods:** Current vaccine equity was investigated with simple linear regression between the county mean HPI and both single and full-dose vaccination rate. Future vaccination coverage per county were predicted using a compartmental mathematical model based on the average rate over the previous 30 days with four different rate-change scenarios.

**Results:** County mean HPI had a strong positive association with both single and full dose vaccination rates (*R*^2^: 0.716 and 0.737, respectively). We predict the overall state rate will exceed 50% fully vaccinated by June 15th if the current rates are maintained; however, the bulk of this coverage comes from the top 18 counties while the remaining 40 counties lag behind.

**Discussion:** The clear association between county HPI and current vaccination rates shows that California is not initiating opening plans from an equitable foundation, despite previous equity programs. If nothing changes, many of the most vulnerable counties will not be prepared to open without consequences come June 15th.

## Introduction

California’s *Blueprint for a Safer Economy Plan* implemented an outbreak severity tier framework for risk management informed by COVID-19 case rates, positive test rates, and hospitalization rates. A health equity revision on October, 2020 required counties with a population above 106,000 (n=36) to ensure that census tracts in the lowest quartile of the Healthy Places Index (HPI), an aggregation of 25 health vulnerability indicators, were not reporting significantly higher positive testing rates than the county average to move to a lower severity tier [1]. Limited vaccination programs began in December 2020, but by March 2021, the CDC listed California’s vaccine distribution as one of the five least equitable programs in the United States [2]. On March 4, 2021 governor Gavin Newsom prioritized 40% of California’s vaccine supply to be administered among the census tracts in the lowest HPI quartile. Updated guidance from the *Blueprint*, which included higher case rate and test positivity thresholds for the most severe tiers, was issued when 2 million and 4 million equity-reserved doses were distributed as an effort to encourage increased testing rates for continued monitoring.

On June 15, 2021, California plans to “fully open” by lifting most business restrictions. Four primary indicators will be used to assess readiness: county-wide adjusted daily case rates and test positivity rates, and both a health and a vaccine equity measurement based on test positivity rates and total vaccinations in the lowest-quartile HPI tracts, respectively. With less than a month to go before widespread protections are lifted, this is a critical time to reassess how equitable California’s vaccine distribution has been. In this short investigation, we examined the current relationship between county-aggregated HPI and vaccination rates, and followed up with simple forecasts of COVID-19 vaccine coverage in California by June 15, 2020 across multiple scenarios to highlight disparities in vaccine uptake across the state.

## Materials and methods

HPI data for individual census tracts were obtained from the Public Health Alliance of California [3]. Single dose and full vaccination rates for each county were obtained from the California Department of Public Health [4]. To assess whether broader patterns of inequity still exist, a linear model was constructed in which the county mean HPI 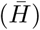 was the predictor for the number of vaccinated individuals per 1,000 population (*Î*) as of May 16th, according to equation 1. Alpine and Sierra counties had extremely small populations and were excluded from this analysis. Regression analysis was executed with R 4.1.0 (R Core Team 2021).

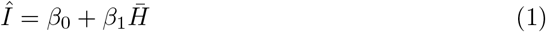

To model future vaccination coverage, we constructed a compartmental model that includes the dynamics between the unvaccinated population, those who got the first vaccine dose and those who are fully vaccinated. Let *W* be the unvaccinated population at time *t*. We assume that no vaccines have been administered at *t* = 0, which implies *W* (0) = *N*. Then, by assuming that we vaccinate individuals at a constant rate for both first dose and second dose proportional to the actual population, we have *W* (*t*), *V*_1_(*t*) and *V*_2_(*t*) that satisfy the following system,

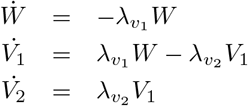

where *W* (0) = *N, V*_1_(0) = 0, and *V*_2_(0) = 0. We stored the cumulative vaccinated population as:

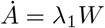

Since real-world vaccination rates have changed since the start of the vaccination, we used the prior 30 days of real-world vaccination data to adjust our model rates. Beginning May 4th, we estimated the growth of single-dose and full vaccination coverage through June 15th for four future scenarios: maintaining the 30-day average rate, a 30% increase in the current average rate, and both a 30% and 60% decrease in the current average rate. Modeling was implemented in python.

## Results

There was a clear, positive association between the county mean HPI and the current proportion of people vaccinated. Regression coefficients were 298.2 (95% CI: 248.1-348.2) for single-dose rate and 248.8 (95% CI: 209.2-288.5) for fully vaccinated rates, with *R*^2^ values of 0.716 and 0.737, respectively. Counties in the San Joaquin Valley region had consistently low values for both HPI and vaccination rates (Figure 1). Compartmental model results are shown in Figure 2. The current trajectory predicts that 66.2% of the population will have at least one dose of the vaccine, and 53.8% will be fully vaccinated. If rates were to increase by 30%, the model predicts that 70.2% of the population will have at least one dose, and 59.0% will be fully vaccinated by June 15th.

**Fig 1.** Simple linear regression of vaccination rates in California counties. *A*: Single dose vaccination rate vs. HPI. *B*: Full dose vaccination rate vs. HPI. Alpine and Sierra counties excluded for small population size.

**Fig 2.** Statewide predictions from the simple compartmental model. Left: Projection for total number of Californians with at least one vaccine dose by June 15th. Right: Projections for total number of Californians who are fully vaccinated by June 15th.

When the model predictions are broken down by county, however, it becomes clear that the state-level results hide some disparities. At current vaccination rates, only 18 out of 58 counties are projected to reach or exceed a fully vaccinated rate of 50%. Even a 30% increase in the current average rate predicts that only 21 out of 58 will reach that threshold (Table 1).

**Table 1.** Percentage of people vaccinated by May 17, 2021 and estimated vaccine coverage by June 15, 2021 in California counties. Vaccine data from [4], updated on May 16, 2021. County population from [5].

| County | Population | Percentage vaccinated |  | Estimated vaccine coverage |  |  |  |
| --- | --- | --- | --- | --- | --- | --- | --- |
| | | ≥ dose | Fully vaccinated | Current rate | 30% reduction | 60% reduction | 30% increase |
| Alpine | 1,129 | 73 | 59 | 81 | 77 | 72 | 85 |
| Marin | 258,826 | 71 | 60 | 73 | 70 | 66 | 76 |
| Santa Clara | 1,927,852 | 64 | 50 | 69 | 64 | 59 | 73 |
| San Francisco | 881,549 | 67 | 53 | 67 | 63 | 58 | 71 |
| Alameda | 1,671,329 | 61 | 49 | 63 | 59 | 55 | 66 |
| Contra Costa | 1,153,526 | 59 | 50 | 62 | 59 | 56 | 65 |
| San Mateo | 766,573 | 65 | 50 | 62 | 58 | 55 | 65 |
| Napa | 137,744 | 63 | 50 | 61 | 57 | 54 | 64 |
| Santa Cruz | 273,213 | 61 | 45 | 61 | 58 | 53 | 65 |
| Sonoma | 494,336 | 60 | 47 | 59 | 56 | 52 | 62 |
| San Diego | 3,338,330 | 62 | 42 | 55 | 51 | 48 | 58 |
| Mono | 14,444 | 55 | 46 | 54 | 52 | 50 | 55 |
| Santa Barbara | 446,499 | 52 | 42 | 54 | 51 | 47 | 56 |
| Ventura | 846,006 | 52 | 41 | 54 | 51 | 47 | 57 |
| Orange | 3,175,692 | 52 | 41 | 53 | 49 | 46 | 56 |
| San Benito | 62,808 | 46 | 35 | 51 | 47 | 43 | 55 |
| Yolo | 220,500 | 53 | 40 | 51 | 48 | 44 | 54 |
| Los Angeles | 10,039,107 | 51 | 39 | 50 | 47 | 43 | 53 |
| Mendocino | 86,749 | 51 | 40 | 49 | 47 | 45 | 52 |
| Placer | 398,329 | 48 | 39 | 49 | 46 | 43 | 51 |
| Inyo | 18,039 | 47 | 40 | 48 | 45 | 43 | 50 |
| Sierra | 3,005 | 46 | 43 | 48 | 47 | 46 | 49 |
| Humboldt | 135,558 | 47 | 38 | 47 | 45 | 42 | 49 |
| Monterey | 434,061 | 49 | 38 | 47 | 44 | 40 | 49 |
| San Luis Obispo | 283,111 | 50 | 40 | 47 | 44 | 41 | 50 |
| Sacramento | 1,552,058 | 45 | 35 | 46 | 43 | 40 | 49 |
| El Dorado | 192,843 | 45 | 34 | 45 | 42 | 39 | 47 |
| Plumas | 18,807 | 42 | 36 | 44 | 43 | 41 | 46 |
| Solano | 447,643 | 48 | 35 | 44 | 42 | 39 | 47 |
| Imperial | 181,215 | 44 | 30 | 43 | 40 | 37 | 46 |
| Nevada | 99,755 | 46 | 36 | 42 | 39 | 37 | 44 |
| San Joaquin | 762,148 | 40 | 28 | 41 | 38 | 35 | 43 |
| Colusa | 21,547 | 38 | 27 | 40 | 37 | 34 | 42 |
| Amador | 39,752 | 46 | 30 | 37 | 35 | 33 | 39 |
| Sutter | 96,971 | 39 | 31 | 37 | 35 | 33 | 39 |
| Tuolumne | 54,478 | 41 | 34 | 37 | 36 | 34 | 39 |
| Calaveras | 45,905 | 41 | 32 | 36 | 34 | 32 | 38 |
| Fresno | 999,101 | 41 | 31 | 36 | 34 | 33 | 38 |
| Lake | 64,386 | 39 | 32 | 36 | 35 | 33 | 38 |
| Riverside | 2,470,546 | 40 | 31 | 36 | 34 | 31 | 38 |
| Stanislaus | 550,660 | 41 | 29 | 36 | 34 | 32 | 39 |
| Butte | 219,186 | 38 | 32 | 35 | 34 | 32 | 37 |
| Glenn | 28,393 | 36 | 30 | 35 | 33 | 31 | 37 |
| San Bernardino | 2,180,085 | 37 | 29 | 35 | 32 | 30 | 37 |
| Mariposa | 17,203 | 41 | 24 | 33 | 30 | 28 | 35 |
| Madera | 157,327 | 37 | 29 | 32 | 30 | 29 | 34 |
| Tulare | 466,195 | 35 | 28 | 32 | 30 | 29 | 33 |
| Siskiyou | 43,539 | 36 | 28 | 31 | 30 | 29 | 32 |
| Trinity | 12,285 | 35 | 27 | 31 | 29 | 28 | 32 |
| Kern | 900,202 | 34 | 27 | 30 | 28 | 26 | 31 |
| Merced | 277,680 | 33 | 22 | 30 | 28 | 26 | 32 |
| Modoc | 8,841 | 30 | 27 | 29 | 28 | 27 | 30 |
| Shasta | 180,080 | 34 | 26 | 29 | 27 | 26 | 30 |
| Del Norte | 27,812 | 33 | 27 | 28 | 26 | 25 | 29 |
| Yuba | 78,668 | 28 | 22 | 27 | 26 | 24 | 29 |
| Kings | 152,940 | 27 | 20 | 23 | 22 | 21 | 24 |
| Tehama | 65,084 | 25 | 21 | 23 | 22 | 21 | 24 |
| Lassen | 30,573 | 22 | 19 | 21 | 20 | 19 | 21 |

## Discussion

When observed in the aggregate as an entire state, it is tempting to say that California is in an optimistic position for a general easing of restrictions come June 2021. However, the successes in large, urban centers have not been similarly realized across the state. As of the writing of this paper, there is a strong correlation between the average HPI of a county and current vaccination rates. The counties that are least vaccinated and least likely to reach acceptable immunization thresholds by June 15th are also those with the most vulnerable populations.

Hesitancy is frequently cited as a stumbling block for achieving high vaccination rates. Vaccine hesitancy is complex and often driven by social and demographic determinants that evolve over time. The temporary pause of the Janssen COVID-19 vaccine on April 13, 2021, for instance, changed the minds of 39% unvaccinated Hispanic women according to a Kaiser Family Foundation vaccine hesitancy survey. In California, the most vaccine-hesitant counties are estimated to have 16.1% of residents responding as “hesitant or unsure” [6]. Few counties are projected to reach the point where more unvaccinated people are hesitant or unsure than not, leading us to believe that the challenge remains one of inequitable distribution rather than differences in vaccine uptake.

The San Joaquin Valley region stands out as particularly unprepared. Of the eight counties (San Joaquin, Stanislaus, Merced, Madera, Fresno, Kings, Tulare, and Kern), seven are in the lowest quartile for county mean HPI, and Fresno currently leads the fully vaccinated rate among them at 31%. These counties are home to 3.97 million people, which is more than one-tenth of the population of California and a greater total population than 23 other states. The San Joaquin Valley is home to large numbers of Latino agricultural workers, who face systemic socioeconomic and environmental inequities, and who have been shown to be at high risk from COVID-19 [7] [8]. Low vaccination rates and a highly vulnerable population are a recipe for continued outbreaks of COVID-19 well beyond June. This is not limited to the San Joaquin Valley, either; the majority of counties were predicted to lag significantly behind the state rate.

This simple study’s use of an aggregate vulnerability index likely obscures how different aspects of vulnerability have lead to inequities across the state. Closer, more sophisticated examinations are warranted to characterize the factors that lead to the larger-scale inequities described here. What lessons can we learn for future pandemics? What infrastructure and trust are we building now that should be nourished beyond COVID-19 to enable more effective and equitable interventions for daily life and the next health crisis?

## Data Availability

Model code is available at our persistent repository (10.5281/zenodo.4795419).

https://github.com/yurygarcia26/California_Vaccine_Coverage/tree/V1

## Acknowledgements

The authors would like to thank the California Department of Public Health and the Public Health Alliance of Southern California for making and maintaining free, public sources of health data.

